# Serological Analysis Reveals an Imbalanced IgG Subclass Composition Associated with COVID-19 Disease Severity

**DOI:** 10.1101/2020.10.07.20208603

**Authors:** Jennifer L. Yates, Dylan J. Ehrbar, Danielle T. Hunt, Roxanne C. Girardin, Alan Dupuis, Anne F. Payne, Mycroft Sowizral, Scott Varney, Karen E. Kulas, Valerie L. Demarest, Kelly M. Howard, Kyle Carson, Margaux Hales, Monir Ejemel, Qi Li, Yang Wang, Nicholas J. Mantis, Kathleen A. McDonough, William T. Lee

**Affiliations:** Division of Infectious Diseases, Wadsworth Center, New York State Department of Health, Albany, NY, 12208 USA; MassBiologics of the University of Massachusetts Medical School, Boston, MA, 02126 USA; Department of Surgery Albany Medical College, Albany, NY,12208 USA; Biomedical Sciences, The School of Public Health, The University at Albany, Albany, NY, 12222 USA

## Abstract

COVID-19 is associated with a wide spectrum of disease severity, ranging from asymptomatic to acute respiratory distress syndrome (ARDS). Paradoxically, a direct relationship has been suggested between COVID-19 disease severity, and the levels of circulating SARS-CoV-2-specific antibodies, including virus neutralizing titers. Through a serological analysis of serum samples from 536 convalescent healthcare workers, we found that SARS-CoV-2-specific and virus-neutralizing antibody levels were indeed elevated in individuals that experienced severe disease. The severity-associated increase in SARS-CoV-2-specific antibody was dominated by IgG, with an IgG subclass ratio skewed towards elevated receptor binding domain (RBD)- and S1-specific IgG3. However, RBD- and S1-specific IgG1, rather than IgG3 were best correlated with virus-neutralizing titers. **We propose that Spike-specific IgG3 subclass utilization contributes to COVID-19 disease severity through potent Fc-mediated effector functions**. These results have significant implications for SARS-CoV-2 vaccine design, and convalescent plasma therapy.

## Introduction

The novel coronavirus, severe acute respiratory syndrome coronavirus-2 (SARS-CoV-2) is the causative agent of COVID-19, a disease responsible for more 1 million deaths in just a matter of months. The case incidence based on virus detection estimates 35 million cases globally to date. However, if population-based serological surveys of SARS-CoV-2 are taken into account, the infection rate of SARS-CoV-2 is likely much higher (McLaughlin et al., 2020; Pollan et al., 2020). This discrepancy highlights the variability of COVID-19 disease presentation in the human population. The severity of disease caused by SARS-CoV-2 infection ranges from an asymptomatic presentation, to acute respiratory distress syndrome (ARDS), and death. Risk factors such as age, gender, and underlying disease are known to be associated with COVID-19 disease severity; however, a subset of patients with severe disease are younger without obvious comorbidities. The immunological features associated with severe COVID-19 disease include high levels of inflammatory cytokines, low lymphocyte counts, high neutrophil to lymphocyte ratios, and increased serum proteins such as C-reactive protein (CRP), ferritin, and D-dimer (Chen et al., 2020a; Giamarellos-Bourboulis et al., 2020; Kuri-Cervantes et al., 2020; Mathew et al., 2020). In addition, several studies have shown that SARS-CoV-2-specific antibody and neutralizing titers are increased in patients who exhibit more severe disease (Long et al., 2020; Piccoli et al., 2020). Therefore, it is important to consider that SARS-CoV-2 specific antibodies may play multiple roles in COVID-19 pathogenesis, including control of viral infection, disease resolution, and immunopathology.

The humoral response to SARS-CoV-2 is primarily directed towards the nucleocapsid (N) protein, and the spike protein that decorates the surface of the virus. The N protein is an RNA binding protein composed of an N-terminal RNA binding domain, and a C-terminal oligomerization domain that are essential for viral RNA transcription and replication (Kang et al., 2020). The spike protein is a multi-domain trimeric glycoprotein composed of two distinct subunits. The S1 subunit is composed of four domains, including the receptor binding domain (RBD) of the spike glycoprotein (Wrapp et al., 2020). The S2 domain forms the stalk-like portion of the full-length trimeric protein and is responsible for viral fusion with the host cell membrane. Antibody responses directed at the spike protein and RBD in particular, have been identified as the main neutralizing component of the SARS-CoV-2 antibody response (Chen et al., 2020b; Robbiani et al., 2020; Suthar et al., 2020). A recent study found that distinct antibody signatures could be linked to different COVID-19 disease outcomes. Specifically, early spike-specific responses were associated with a positive outcome (convalescence), while early N-specific responses were associated with a negative outcome (death). Moreover, the Fc-associated functions of the antibody response such as antibody-mediated phagocytosis, cytotoxicity, and complement deposition were critical for disease resolution (Atyeo et al., 2020). However, little is known about antibody isotypes and subclasses generated in response to SARS-CoV-2, or their role in COVID-19 pathogenesis. In this study, we analyzed the humoral immune response to SARS-CoV-2 in a unique cohort of 536 convalescent healthcare workers that were stratified by COVID-19 disease severity. This cohort provided us with a unique snapshot of the SARS-CoV-2 specific antibody profile at convalescence, as a window to previous disease pathogenesis. We found a significantly increased SARS-CoV-2 specific antibody response in severe COVID-19 patients when compared to patients who experienced mild and moderate disease symptoms. This severity-associated antibody increase was dominated by IgG, with a disproportionate IgG subclass response dominated by IgG3.

## Results

### Study Cohort

A total of 536 COVID-19 serum specimens from convalescent healthcare workers (HCW) were received by the Wadsworth Center for SARS-CoV-2 serology testing. The sera were obtained from individuals who had tested positive by RT-PCR and had illness consistent with SARS-CoV-2 infection. Table 1 provides basic patient demographic information stratified by self-reported COVID-19 disease severity. The gender distribution of the study cohort was bias towards females (29% male, 69% female, with 2% gender unknown) reflecting the gender disparity within the HCW. The mean age and days post-onset of symptoms (DPO) was roughly the same in all gender categories. Approximately 10% of the study cohort experienced severe disease, 40% moderate disease, 40% mild disease, with 10% uncharacterized disease. The average age of each group in our cohort increased with disease severity, as did the percentage of males within each group. In fact, the representation of males (65%) in the severe group was more than double that of the mild group (31%), illustrating a clear gender bias in COVID-19 disease severity.

**Table 1:** Study Cohort of Convalescent Healthcare Workers.

|  | **All** | **Male** | **Female** | **Unknown** |
| --- | --- | --- | --- | --- |
| **Count** | 536 | 154 (29%) | 370 (69%) | 12 (2%) |
| **Mean Age (years)** | 40 | 40 | 40 | 40 |
| **Days Post Onset** | 41 | 40 | 41 | 42 |
| **% Mild Disease** | 40.5% | 44.2% | 38.9% | 41.7% |
| **% Moderate Disease** | 40.1% | 35.0% | 42.1% | 41.7% |
| **% Severe Disease** | 9.1% | 10.4% | 8.7% | 8.3% |
| **% Unknown Severity** | 10.3% | 10.4% | 10.3% | 8.3% |

### Relationship of Antibody Production and Virus Neutralization Capability with COVID-19 Disease Severity

A total of 536 serum samples were assessed for SARS-CoV-2-specific antibodies using a clinical microsphere immunoassay (MIA) to detect total antibody directed against N protein or RBD of SARS-CoV-2. We used the median fluorescence intensity (MFI) of PE-labeled anti-human Ig to antigen-coupled beads as a qualitative measure of SARS-CoV-2-specific antibody abundance in donor serum. We found that total serum antibody specific to the N protein, and RBD of SARS-CoV-2 were increased with increasing disease severity (**Table 1; Figure 1a**). To evaluate the relationship between overall antibody levels and protective antibody we measured viral neutralization using a plaque reduction neutralization test (PRNT) where the highest dilution of sera providing 50% (PRNT50) or 90% (PRNT90) viral plaque reduction relative to a virus only control was reported as the neutralizing titer. Both PRNT50 and PRNT90 measurements identified a concomitant increase in virus neutralizing titers with disease severity **(Figure 1b**). RBD-specific responses were strongly correlated with virus neutralization, as revealed through a Spearman’s correlation analysis with PRNT90 values and MFI values. The correlation with PRNT90 titers was greater for RBD-specific Ig than N-specific Ig (r = 0.68 and 0.53, respectively)(**Figure 1c**). Since the function of the RBD is host cell attachment through ACE-2 binding, our data suggests that the neutralization activity of SARS-CoV-2-specific antibodies in our PRNT assay is primarily based on the ability to block viral attachment and uptake by host cells (Chen et al., 2020b; Robbiani et al., 2020; Suthar et al., 2020).

**Figure 1:**
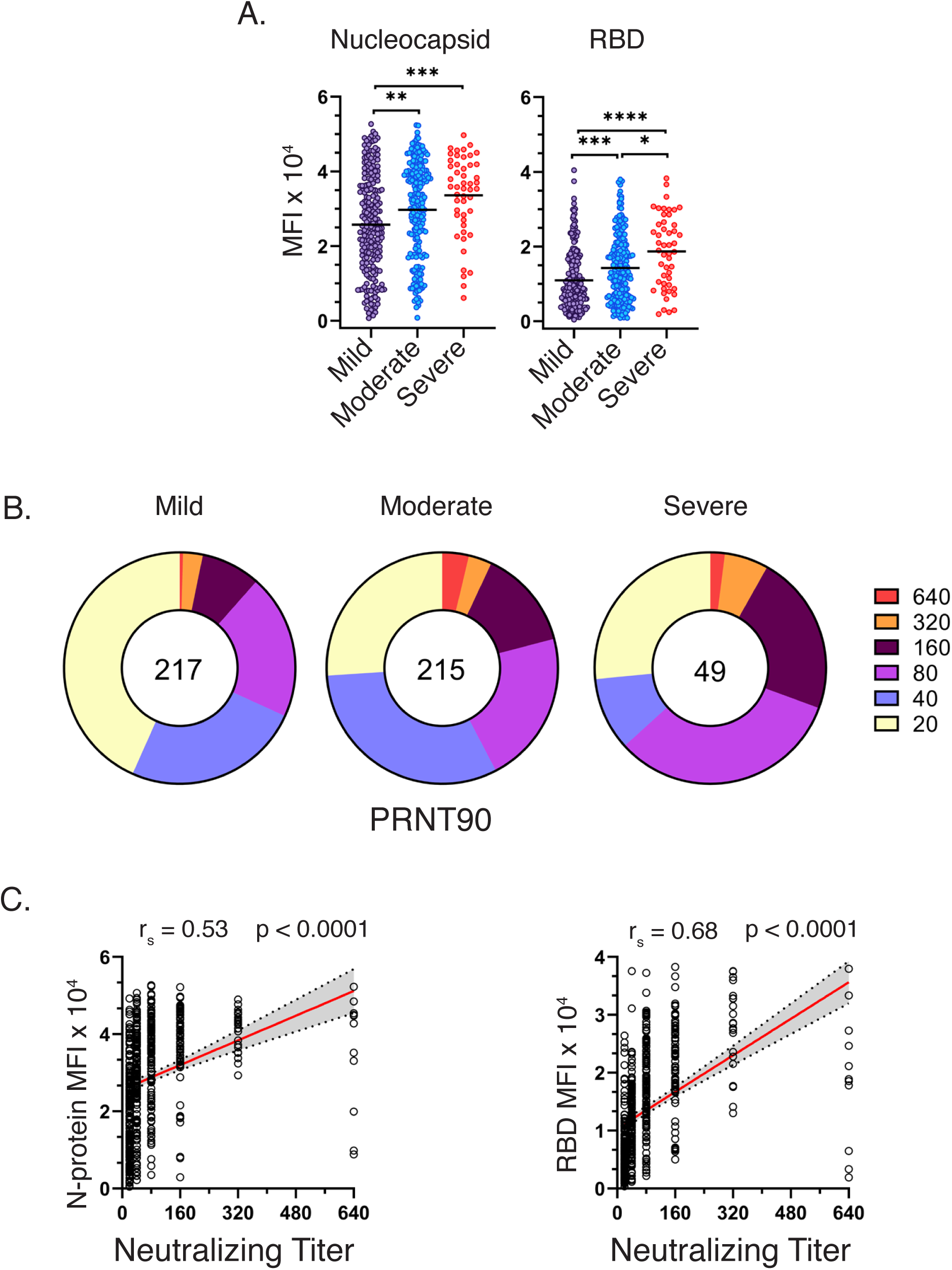
Relationship of Antibody Production and Virus Neutralization Capability with COVID-19 Disease Severity. **(a)** Total Ig reactivity (MFI) to SARS-CoV-2 nucleocapsid and RBD from convalescent COVID-19 individuals as grouped by disease severity. **(b)** Spearman’s correlation of the nucleocapsid and RBD MFI from the full cohort of 536 patients. **(c)** Reciprocal PRNT90 dilutions from the same convalescent COVID-19 individuals, grouped by disease severity. **(d)** Spearman’s correlation of PRNT90 dilutions versus the nucleocapsid or RBD MFI from the full cohort of convalescent individuals.

### Isotype and Antigen Distribution of the SARS-CoV-2 specific Antibody Profile Across COVID-19 Disease Severities

To better resolve the SARS-CoV-2 humoral immune response across COVID-19 severity groups, we tested additional antibody isotypes and specificities in our clinical MIA assay. Positive antibody reactivity was defined as 6 standard deviations above the mean MFI, as determined by a panel of 93 pre-COVID-19 normal human serum specimens tested against each antibody isotype/antigen combination. Index value measurements were calculated by dividing the MFI by the previously determined cutoff MFI. In addition to N and RBD, we included the S1 and S2 sub-unit domains as antibody targets allowing us to incorporate all the potential epitopes from the Spike protein, and to assess the contribution of each domain to the overall antibody response. In general, we found that antibody reactivity to SARS-CoV-2 antigens differed dramatically between isotypes. As expected, IgG was the dominant isotype generated in response to SARS-CoV-2 infection, with antibodies from 97-98% of serum specimens yielding positive recognition of all antigens and antigenic subunits tested (**Figure 2a, Table S1**,). IgM and IgA were primarily reactive towards the Spike S1 subunit, including the RBD—with 73% and 79% positivity, respectively. The presence of antigen-specific IgM at this convalescent time-point is notable (∼ day 40), as IgM is generally considered a biomarker of acute-phase infection. IgM and IgA with specificity to the nucleocapsid were rare, with only 22% of specimens testing positive for each isotype, respectively. IgM antibodies with specificity for the Spike S2 subunit were even more rare (5%), while IgA responses displayed a 36% positivity rate. As shown in Figure 1, we observed that total SARS-CoV-2-specific antibody increased with disease severity. We next examined individual isotype distributions across disease categories. There was a >1.5 fold increase in IgG generated in the severe COVID-19 group as compared to mild, against all antigens tested (N, RBD, S1, and S2). In addition, IgM against the N-protein had a 1.6-fold increase in the severe group, as compared to mild, with little or no change in other antigens (**Figure 2b)**. Finally, IgA generated against the N-protein and Spike S1 subunit resulted in a 2.4 and 2.0-fold increase, respectively, the largest fold changes observed between mild and severe disease categories. The increase in N-specific IgA is consistent with the observation that exceptionally high levels of serum IgA were associated with acute respiratory distress syndrome (ARDS)(https://doi.org/10.1101/2020.05.21.108308). Furthermore, the observed correlation of IgM and IgA specific for the N-protein increasing with disease severity is reminiscent of the early N-dominated response in deceased individuals reported by (Atyeo et al., 2020). Our results illustrate that while total SARS-CoV-2 specific antibody levels increase with COVID-19 severity, this change is not equal among antibody specificities or isotypes. The increase in total N-specific antibody and spike-specific IgG suggests there are enhanced levels of viral antigens and corresponding increases in T cell-dependent B cell responses during severe COVID-19.

**Figure 2:**
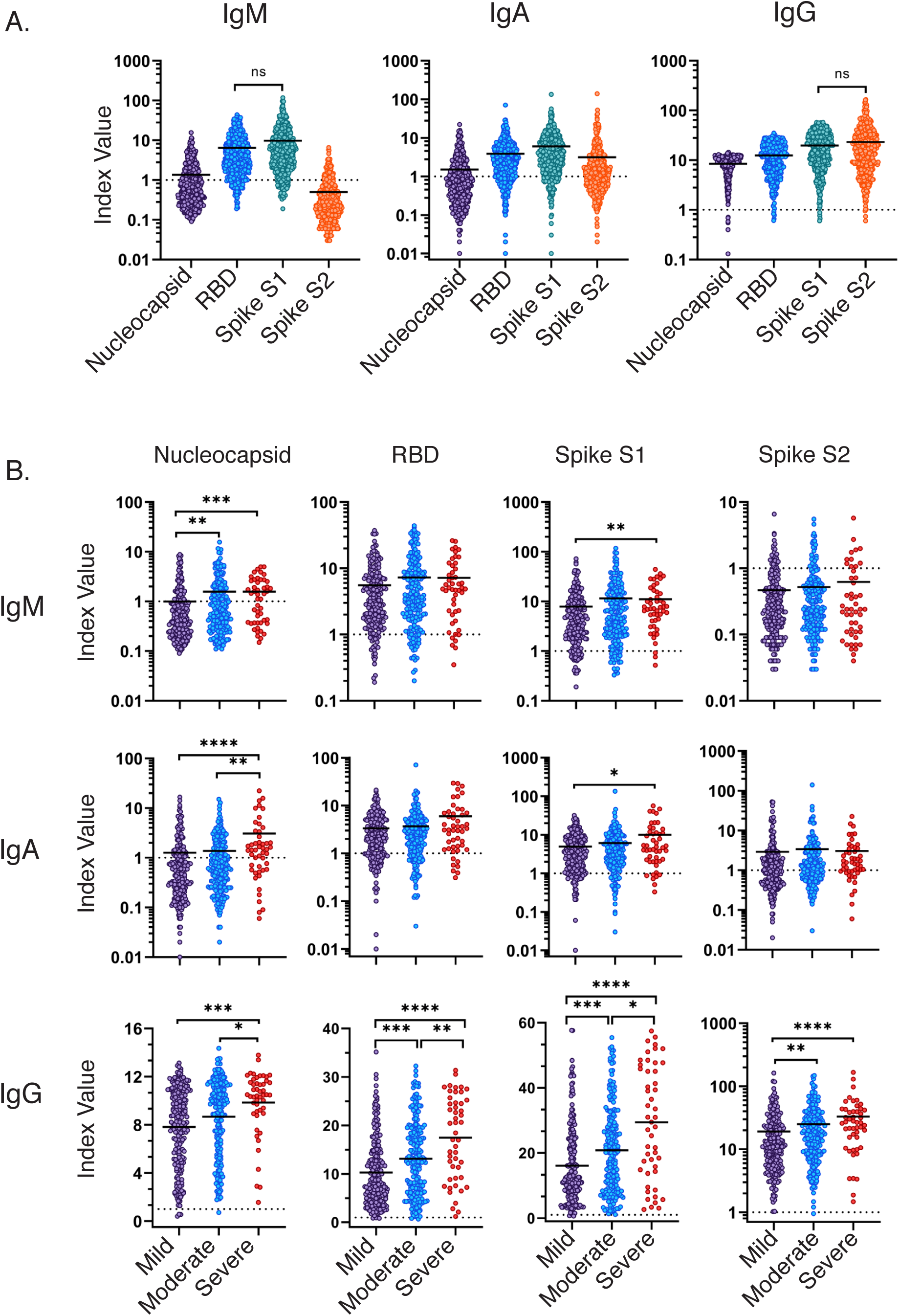
Isotype and Antigen Distribution of the SARS-CoV-2 specific Antibody Response. Reactivity of IgM, IgA, and IgG specific to the SARS-CoV-2 nucleocapsid, RBD, S1 subunit, or S2 subunit of the full patient cohort **(a**) or grouped by disease severity **(b)**. Index value represents the raw MFI divided by the background cutoff value determined by a panel of 93 normal human serum specimens. Statistical significance was determined by the non-parametric Kruskall-Wallace test where * p ≥ 0.05 ** p ≥ 0.001 *** p ≥ 0.0001, and **** p ≥ 0.00001.

### IgG Subclass and Antigen Distribution of the SARS-CoV-2 specific Antibody Profile Across COVID-19 Disease Severities

IgG is the classic antibody isotype involved in T cell-dependent B cell responses, resulting in durable humoral memory. In humans, IgG can be further divided into four functional subclasses – IgG1, IgG2, IgG3, and IgG4. Each subclass has unique properties and effector functions that are primarily driven by the Fc portion of the antibody molecule including complement activation, Fc receptor (FcR) binding, and serum half-life. In particular, the ability of an antibody to bind and signal through FcR on effector cells can have profound effects on disease resolution in many models of infectious disease. Therefore, we sought to characterize the IgG subclass usage of the SARS-CoV-2 antibody response. We found that IgG1 and IgG3 were the dominant IgG subclasses produced in response to SARS-CoV-2 infection (**Figure 3A)**. The responses to nucleocapsid, RBD, and S1 subunit were each dominated by IgG1 with over 90% of specimens testing positive (**Table S2**). Strikingly, the IgG response toward the S2 subunit was dominated by the IgG3 subclass with 94% of specimens yielding a positive result. IgG2 responses were moderate, with a positivity rate ranging from 8 to 21%. Finally, IgG4 responses were especially rare with a positivity rate of 0 – 9% **(Figure S1, Table S2)**. Next, we asked whether the IgG subclass representation changes in relation to COVID-19 disease severity. We observed significant increases in both IgG1 and IgG3 with increasing COVID-19 severity, specific for all antigens tested (N, RBD, S1, and S2) (**Figure 3B**). A small, yet significant increase in N-specific IgG2 was associated with disease severity, while RBD-specific IgG2 trended downward with disease severity. The largest difference was seen with the IgG3-specific response toward the RBD and S1 subunit with a 7 and 6-fold increases in the mean index ratio between the mild and severe groups, respectively. Together, these results highlight distinct differences in the SARS-CoV-2 specific antibody profile among individuals that experience mild, moderate, or severe symptoms of COVID-19.

**Figure 3:**
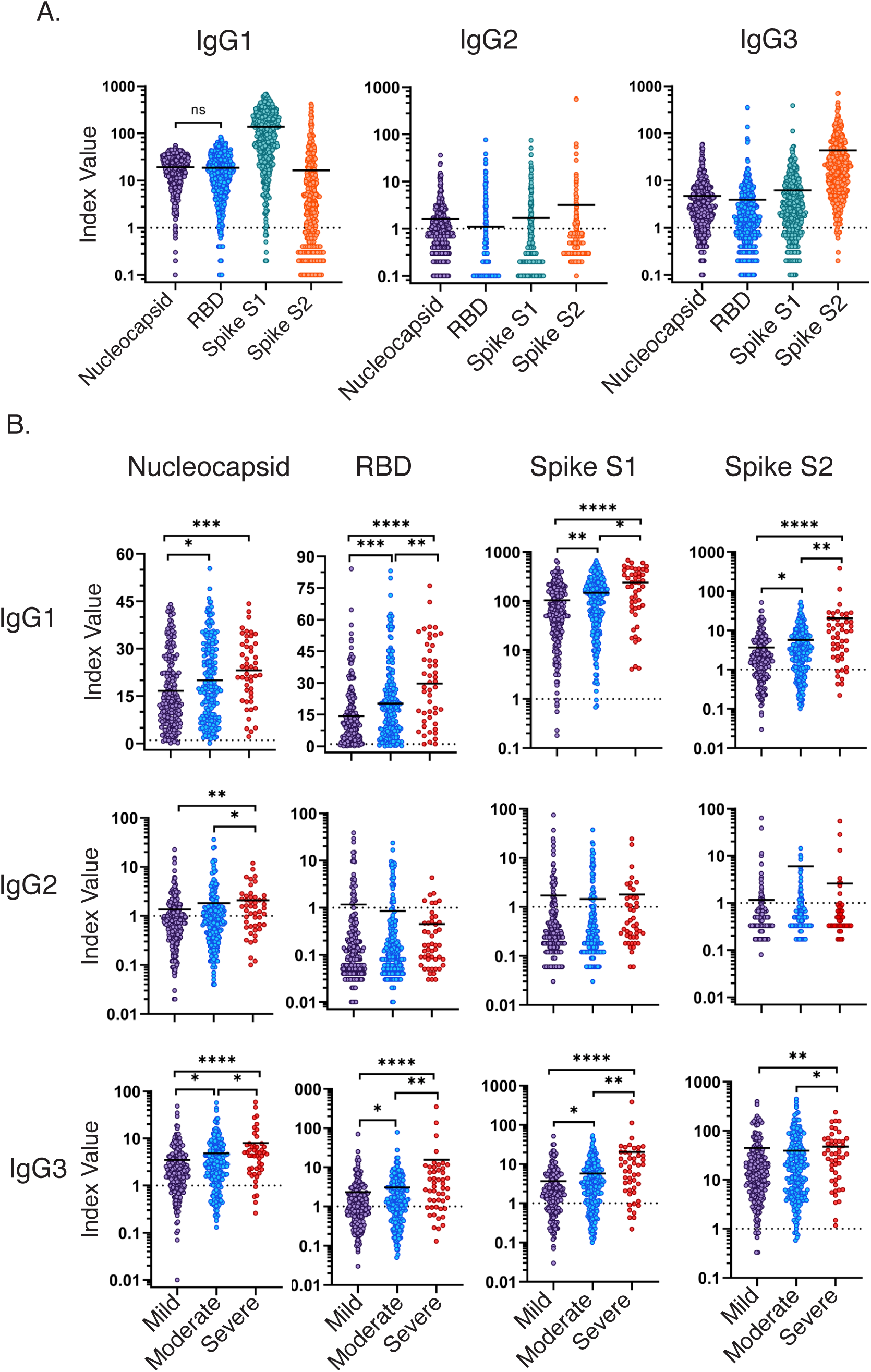
IgG Subclass and Antigen Distribution of the SARS-CoV-2 SARS-CoV-2 specific Antibody Profile Across COVID-19 Disease Severities. **(a)** Reactivity of IgG1, IgG2, and IgG3 to SARS-CoV-2 nucleocapsid, RBD, S1 subunit, or S2 subunit of the full patient cohort **(b)** or grouped by disease severity. Index value represents the raw MFI divided by the background cutoff value determined by a panel of 93 normal human serum specimens. Statistical significance was determined by the non-parametric Kruskall-Wallace test where * p ≤ 0.05 ** p ≤ 0.001 *** p ≤ 0.0001, and **** p ≤ 0.00001.

### Correlation of Antibody Measurements with COVID-19 Disease Severity

We sought to determine a minimal set of criteria that could best distinguish between individuals with different disease severities without overfitting. A training subset of the data was used to create an initial ordered probit regression model that included all potentially predictive variables, including antibody reactivities, sex, age, days post onset, and neutralizing antibody titers. Backwards stepwise selection by Akaike information criterion (AIC) was performed on this model to determine the optimal set of features. When this model’s performance was measured on a testing subset of the data, it displayed higher accuracy (60%, **Table S3**) than both the initial all-inclusive model and any individual univariate model. The combined model that best discriminates between mild, moderate, and severe COVID-19 includes age, RBD-specific IgG1, and S1-specific IgG3. As shown in **Figure 4**, the severe group clusters at high IgG1 RBD, IgG3 S1, and age measurements. In contrast, the mild and moderate groups cluster toward lower IgG1 RBD, IgG3 S1, and age measurements. This model suggests that increased age, and Spike-specific IgG responses play important roles in COVID-19 disease severity. In order to account for the confounding effects of age, a similar series of ordered probit regression models were created that included age as a covariate. All variables that were significantly associated with disease severity in the previous models retained their significant association with disease severity except S2-specific IgG1 (**Table S4**).

**Figure 4:**
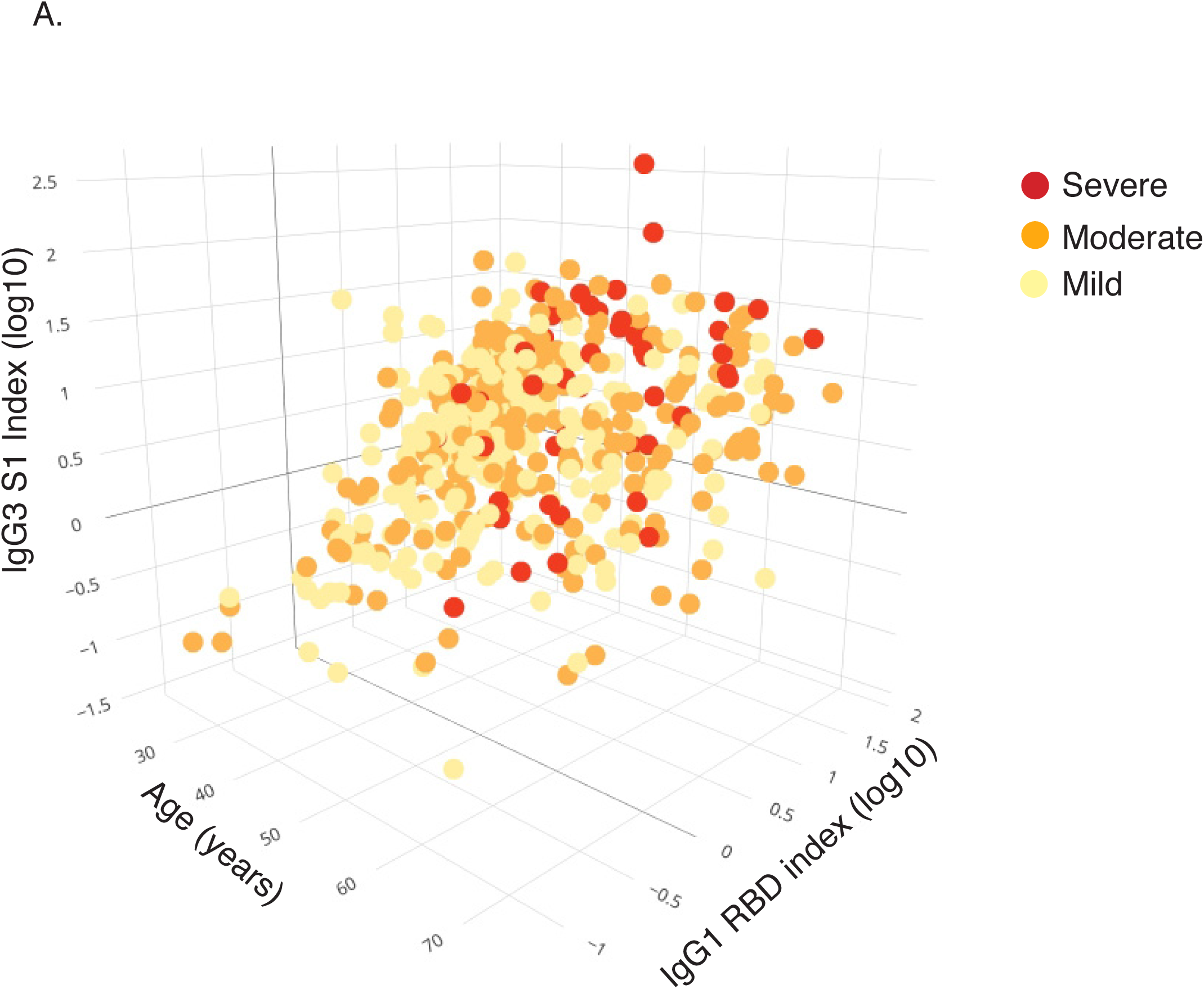
Correlation of Antibody Measurements with COVID-19 Disease Severity. **(a)** Three-dimensional scatter plot depicting the optimal feature set of disease severity-associated features (age, log10-transformed index values (MFI/cutoff) for S1-specific IgG3, and RBD-specific IgG1) as determined by ordered probit regression and backwards stepwise selection by Akaike information criterion. Data is displayed as the distribution of mild (yellow), moderate (orange), and severe (red) disease severities across variables in 478 patients.

### Correlation of IgG Subclasses with COVID-19 Disease Severity

To define the contribution of each IgG subclass to the total SARS-CoV-2 antibody response, we calculated a ratio based on the index value (MFI/cutoff) of each IgG subclass/antigen pair divided by the IgG index value (MFI/cutoff) of the same antigen. While the S1 and RBD-specific IgG1 index ratio remained constant across the severity groupings (**Figure 5A**), there was a significant enrichment of the RBD-specific and S1-specific IgG3 index ratio in severe patients compared to mild (>10-fold and 2-fold increase, respectively). In contrast, a moderate yet significant decrease of the RBD-specific IgG2 index ratio was observed to correlate with severity. To verify the changes in IgG subclass proportions with respect to COVID-19 severity, we evaluated the relative changes in normalized group means across the three severity groups. The relative change between the mild/moderate, and severe groups was most pronounced for RBD-specific IgG2, and RBD/S1-specific IgG3. The group mean MFI of the RBD-specific IgG2 for patients identified with mild or moderate disease (78.3) was higher than the mean for patients identified with severe disease (27.9). Conversely, the RBD/S1-specific mean for patients identified with mild or moderate disease (164.1/142.9) was significantly lower than patients with severe disease (953.6/598.9). A t-test established statistically significant differences between these group means (0.0004, and 0.0483/0.0320, respectively) (**Figure 5B**), reinforcing that divergent IgG subclass responses are associated with COVID-19 disease severity.

**Figure 5:**
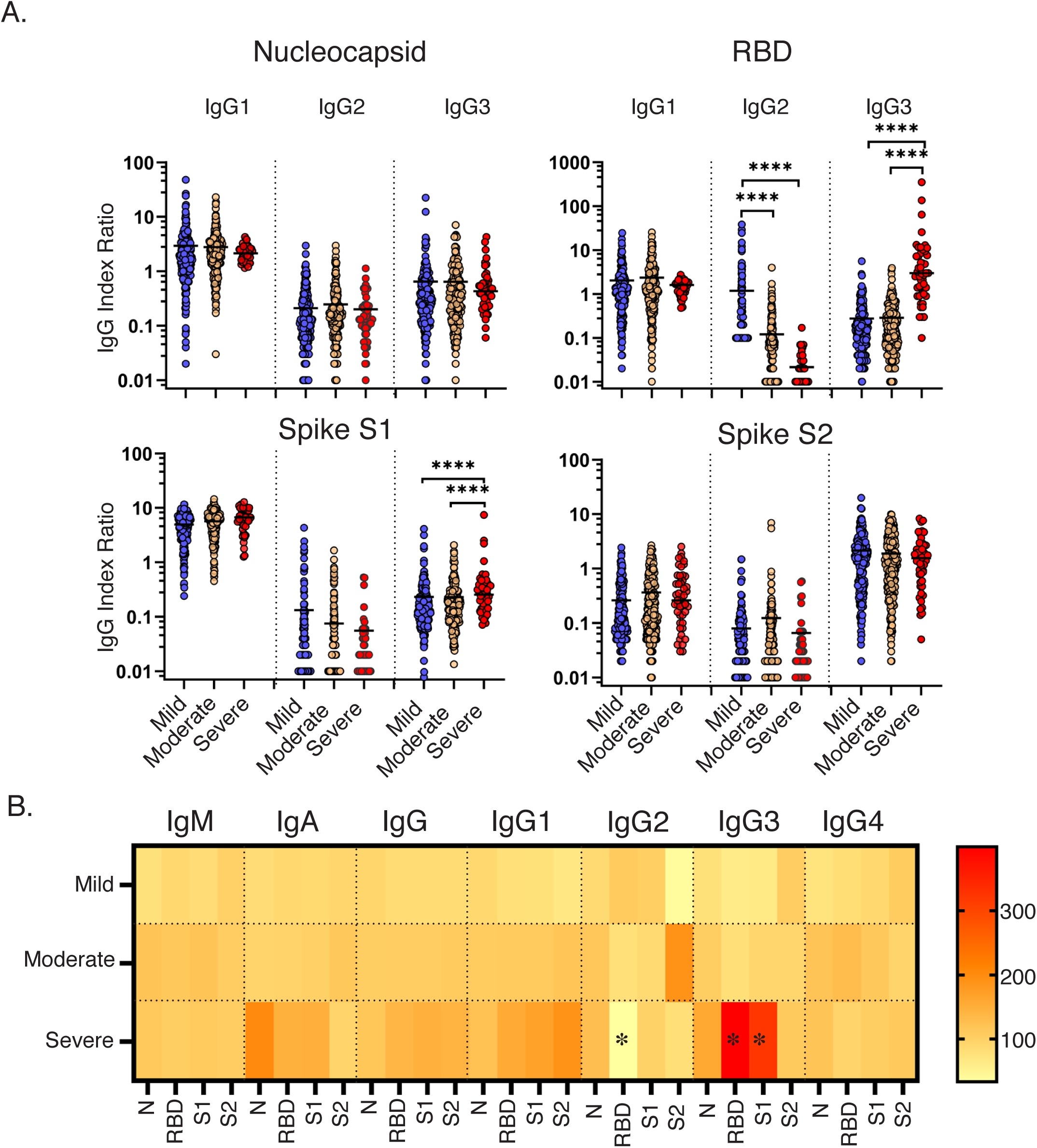
IgG Subclass Ratios Associated with COVID-19 Disease Severity. **(a)** IgG/Subclass index ratios for IgG1, 2, and 3 specific to the nucleocapsid, RBD, S1, and S2 subunits in mild, moderate, and severe patients. **(b)** Heat map represents the average MFI of each severity group divided by the average MFI of the entire data set. * denotes statistical significance. **(c)** Spearman’s correlation coefficient comparing the IgG response to the PRNT90 neutralizing titer.

A correlation network was created to examine the relationship between antibody isotypes, subclasses, antigen specificity, viral neutralization, and clinical features of the HCW dataset. The network was based on a comprehensive correlation matrix (**Figure S2**), and stringently gated on only the strongest associations with a Spearman’s coefficient above 0.65. As expected, S1 and RBD measurements were highly correlated, as were IgG and corresponding IgG subclasses. A dominant network cluster was formed between N-specific IgG, S1-specific IgG (including the RBD domain), and viral neutralizing titers **(Figure 6**). N-specific IgG/IgG1/IgG3 formed its own mini-cluster on the edge of the highly inter-related IgG response to S1 and RBD. In addition, PRNT50 and PRNT90 measurements were very strongly correlated with each other (r_s_ = 0.848), and the PRNT90 measurement was strongly correlated (0.5 ≤ r_s_ ≤ 0.7) with IgG, IgG1, antibodies targeting the S1 domain and RBD of the spike protein. This cluster was also significantly correlated with disease severity, as determined by ordered probit regression (**Table S3**). IgM, IgA, and IgG2 responses to S1 and RBD, were also highly correlated, but separate from the central cluster. Taken together, our results reveal a shift in the proportional Spike-specific IgG response toward the highly inflammatory IgG3 subclass (Vidarsson et al., 2014) that is linked to COVID-19 disease severity, and not with increases in viral neutralizing activity.

**Figure 6:**
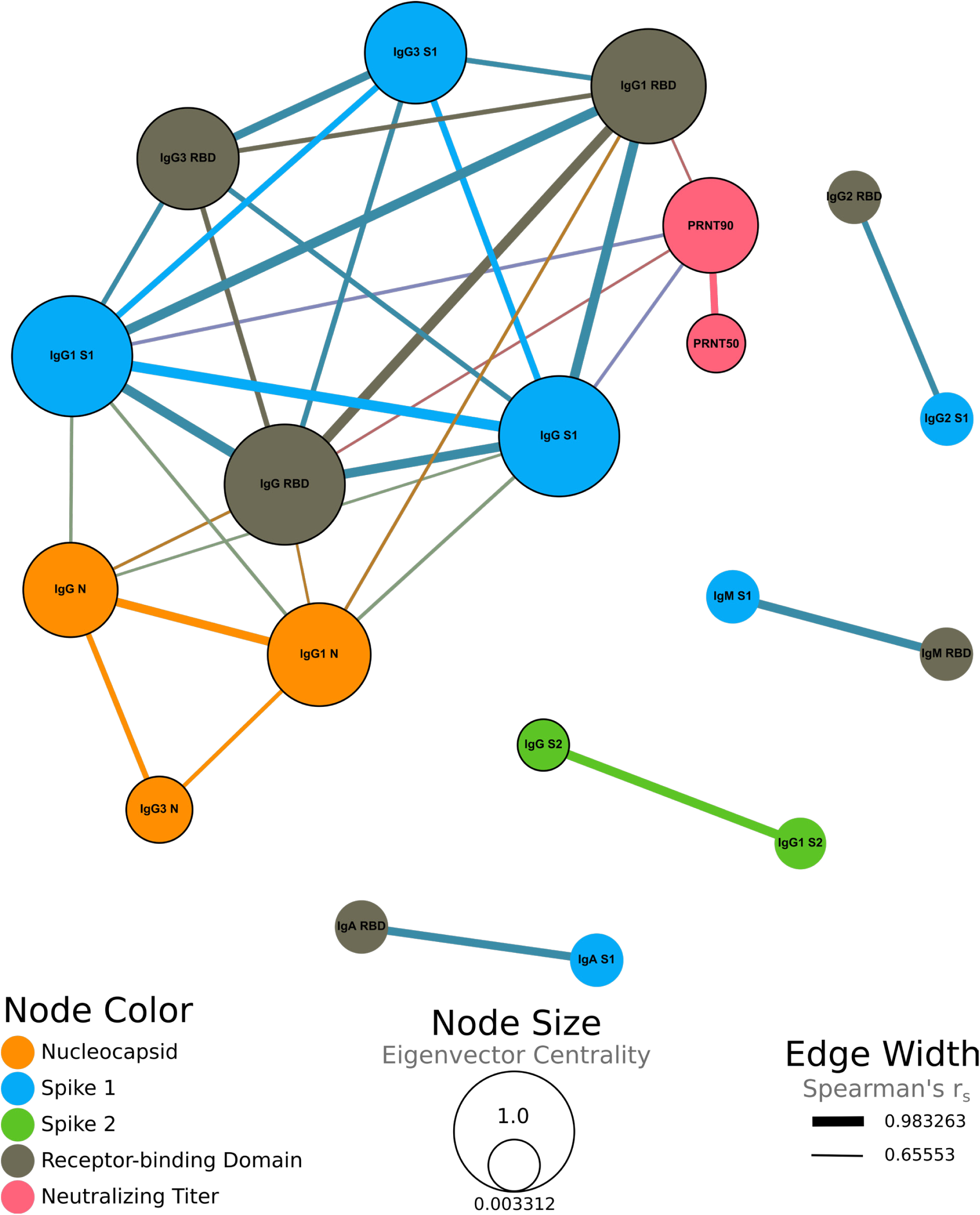
Correlation Network. **(a)** Correlation network displaying strongly correlated (Spearman’s r_s_ > 0.65) variables. Edge thickness represents the magnitude of the correlation between variables. Node size represents eigenvector centrality, showing the influence of each node on the network. Node color represents whether that variable corresponds to an antibody targeting the nucleocapsid (orange), S1 (blue), S2 (green), or RBD (grey) regions, or to neutralizing antibody titers (pink). Black borders around nodes correspond to variables with a significant correlation with severity determined by ordered probit regression modeling, while controlling for age as a confounding variable. All displayed correlations are statistically significant (Benjamini and Hochberg adjusted p < 0.05).

## Discussion

In the present study, we performed a comprehensive analysis of the serum antibody response to SARS-CoV-2 in a cohort of more than 500 recovered HCW who experienced varying degrees of COVID-19 severity. In general, we found that age, total SARS-CoV-2-specific antibody, and viral neutralizing activity was increased in individuals who experienced severe disease. The serological profile for those who experienced severe COVID-19 was characterized by both a global increase in N-specific antibodies, as well as an increase in spike-specific IgG. The overall increase in SARS-CoV-2 specific antibody in severe patients may be due to increased viral loads, and thus increased antigen for driving humoral responses. Paradoxically, high-titer, neutralizing antibodies are known to be protective, and a biomarker of immunity in many viral infections (Ng et al., 2019; Plotkin, 2010). To rectify the disconnect between increased antibodies, increased neutralization, and increased disease severity, we performed a thorough analysis of the IgG subclass response to SARS-CoV-2. Our analysis revealed a substantial difference in the spike-specific IgG subclass composition, where a greater proportion of S1 and RBD-specific IgG3, and lower proportion of S1-specific IgG2 were associated with COVID-19 severity. Spike-specific IgG1, and not IgG3 was most closely correlated with *in vitro* viral neutralization, leading us to the conclusion that excess IgG3 may play an inflammatory role in the pathogenesis of COVID-19 in some individuals.

In viral infections, blockade of cell attachment, and direct viral neutralization are thought to be the primary effector mechanisms through which antibodies contribute to host protection. However, antibodies also facilitate other mechanisms of pathogen clearance through the conserved Fc portion of the molecule. In humans, IgG1 and IgG3 are the most common subclasses elicited by viral infections (Ferrante et al., 1990). While IgG1 is the most abundant subclass in the serum, IgG3 is particularly effective at inducing effector functions through high affinity interactions with complement proteins and by activating type 1 Fc receptors. In fact, IgG3-dependent antibody dependent cellular phagocytosis (ADCP), and cytotoxicity (ADCC), were shown to be critical mechanisms for vaccine-mediated protection against HIV (Chung et al., 2014; Neidich et al., 2019). The importance of antibody-mediated effector functions has also been shown for COVID-19 disease resolution, where Spike-specific ADCP, antibody-dependent neutrophil phagocytosis (ADNP), and antibody-dependent complement deposition (ADCD) were enhanced in COVID-19 survivors. However, Spike-specific antibodies that primarily promoted Fc-mediated NK cell activities were expanded in non-survivors, highlighting the potential for potent Fc-mediated functions to exacerbate pathologic inflammation (Atyeo et al., 2020). The large proportional increase of RBD-specific IgG3 (>10-fold) in severe COVID-19 suggests these antibodies may promote immunopathology rather than tissue repair. Therefore, we propose that an unbalanced IgG response, enriched in IgG3 promotes excess inflammation, exacerbating the symptoms of COVID-19. Supporting our hypothesis, Lui et al. showed that anti-spike IgG promoted acute lung injury in humans and macaques infected with SARS-CoV-1 through macrophage polarization to an inflammatory phenotype (Liu et al., 2019). Serum from COVID-19 patients who experienced severe symptoms was also shown to promote the release of disease promoting neutrophil extracellular traps (NETs), although the stimulating factor was not addressed (Zuo et al., 2020). Finally, both immune complexes and complement pathways were shown to play a pathogenic role in IgG-mediated lung injury in pandemic H1N1 influenza infection (Monsalvo et al., 2011). Future studies are needed to address the FcR dependence and functional differences of IgG1 and IgG3 in COVID-19 serum. Cryoglobulins are a group of antibodies that precipitate in blood at temperatures below 37°C, and their presence is associated with autoimmune and chronic infectious diseases with pathologic effects including vascular, renal, and neurologic complications. Interestingly, IgG3 is the only IgG subclass capable of forming cryoglobulins through Fc-Fc interactions (Otani et al., 2012). Determination of the glycosylation status of SARS-CoV-2 specific antibodies is also warranted to assess the potential inflammatory nature of antibodies in severe COVID-19 (Wang and Ravetch, 2019). In conclusion, we recommend that the contribution of S1 and RBD-specific IgG3 to COVID-19 disease severity should be a strong consideration for both SARS-CoV-2 vaccine design, monoclonal antibody therapeutics, and convalescent plasma therapy.

### Limitations of Study

The size of our patient cohort enabled a robust analysis of the antibody response to SARS-CoV-2 in individuals with self-reported mild, moderate, or severe disease. However, the addition of samples from the of the full spectrum of COVID-19 presentation, including asymptomatic individuals, and non-survivors would significantly strengthen our study. The convalescent time-point at which this study was conducted allowed us to capture a large cohort that was with well documented clinical criteria.

However, the late time-point (day 40 post onset) of this study weakens the predictive capacity of our analysis for earlier points during infection. In future studies we would like to follow a comprehensive cohort of individuals from COVID-19 symptom onset through and beyond recovery to assess how the early immune response influences both disease severity, and durable memory.

## Data Availability

Data used in the manuscript is not publicly available.

## Acknowledgements

We acknowledge and thank the members of the following Wadsworth Center laboratories and core facilities for their expert technical assistance: Diagnostic Immunology, Steven Bush, Andrea Furuya, Theresa Lamson, Mary Marchewka, Randy Stone, Colleen Walsh, and Casey Warszycki; and the Tissue Culture Core. We thank Elizabeth Leadbetter and Gary Winslow for providing helpful discussions and critical reading of the manuscript.

## Author Contributions

J.Y. Designed experiments, conducted experiments, analyzed and interpreted data, and wrote the paper.

D.E. Designed statistical models, performed statistical analyses, assisted with data interpretation, and created figures.

D.H. Designed experiments, conducted experiments, and assisted in data analysis.

R.G., A.D., and A.P. designed and performed PRNT assays.

M.S. and S.V. Performed statistical analysis.

K.K., V.D., K.H., K.C, and M.H. assisted with data entry and running experiments.

M.E., Q.L., and Y.W. were responsible for generating the RBD and S1 subunit proteins used in the MIA.

N.M., K.M., and W.L. Oversaw the design, implementation, and analysis of experiments/data.

## Materials and Methods

### COVID-19 Serum Samples

Studies were performed on sera from clinical specimens submitted to the Wadsworth Center, New York State Department of Health for determination of antibody reactivity to SARS-CoV-2. The study population were recovered individuals who were all RT-PCR confirmed by Roche COVAS 6800 or Cepheid XPert for the presence of SARS-CoV-2 and who had a self-reported degree of disease severity (mild, moderate, or severe). All individuals had recovered from the disease and had defined days between the onset of symptoms and sample collection. Specimens were stored at 4°C until clinical tested was completed (>1 week) and transferred to −80°C for long-term storage. Aliquots were made to minimize freeze-thaw. All testing and archiving of human specimens was approved by NYSDOH Institutional Review Board (IRB 20-021).

### Reagents

For MIAs, wash buffer and phosphate buffered saline pH 7.4, 0.05% sodium azide (PBS-TN) were purchased from Sigma-Aldrich (St. Louis, MO). Chemicals, 1-ethyl-3-(3-dimethylaminopropyl) carbodiimide hydrochloride (EDC) and N-hydroxysulfosuccinimide (sulfo-NHS), were supplied by Pierce Chemicals (Pierce, Rockford, IL). Microspheres, calibration microspheres, and sheath fluid were obtained from Luminex Corporation (Luminex Corp., Austin, TX). R-phycoerythrin (PE)-conjugated goat anti-human Ig, goat anti-human IgG, anti-human IgM, anti-human IgA, mouse anti-human IgG1, mouse anti-human IgG2, mouse anti-human IgG3, and mouse anti-human IgG4 were purchased (Southern Biotech). Recombinant SARS-CoV-2 nucleocapsid and spike S2 domain were purchased (Native Antigen, Oxfordshire, UK). Recombinant SARS-CoV-2 RBD and spike S1 domain were provided by MassBiologics (Boston, MA), and produced as described below.

### Spike Glycoprotein Expression and Purification

The amino-acid sequence of the SARS-CoV-2 S glycoprotein sequence (GeneBank: MN908947) were used to design a codon-optimized version for mammalian cell expression. The synthetic gene encoding the Receptor Binding Domain (RBD) a.a. 319-541 and S1 subunit a.a. 1-604 of the S glycoproteins were cloned into pcDNA 3.1 Myc/His in-frame with c-Myc and 6-histidine epitope tags that enabled detection and purification. The cloned genes were sequenced to confirm that no errors had accumulated during the cloning process. All constructs were transfected into Expi293 cells using ExpiFectamine 293 Transfection Kit (Thermo Fisher). And recombinant proteins were purified by immobilized metal chelate affinity chromatography using nickel-nitrilotriacetic acid (Ni-NTA) agarose beads. Proteins were eluted from the columns using 250 mmol/L imidazole and then dialyzed into phosphate-buffered saline (PBS), pH 7.2 and checked for size and purity by sodium dodecyl sulfate polyacrylamide gel electrophoresis (SDS-PAGE).

### Microsphere Immunoassay

Specimens were assessed for the presence of antibodies reactive with SARS-CoV-2 using an MIA modified from a previously described procedure (Wong et al., 2017). Briefly, recombinant SARS-CoV-2 nucleocapsid and spike RBD, S1, and S2 subunits were covalently linked to the surface of fluorescent microspheres (Luminex Corporation). Serum samples (25 µL at 1:100 dilution) and antigen-conjugated microspheres (25 µL at 5⨯10^4^ microspheres/mL) were mixed and incubated 30 minutes at 37°C before washing and further incubation with phycoerythrin (PE)-conjugated secondary antibody. The PE-conjugated antibodies were chosen to specifically recognize, as indicated, total antibodies (pan-Ig), or, individually IgM, IgA, IgG, IgG1, IgG2, IgG3, IgG4. After washing and final resuspension in buffer, the samples were analyzed on a FlexMap 3D analyzer using xPONENT software, version 4.3 (Luminex Corporation).

### Plaque Reduction Neutralization Assay (PRNT)

This assay has been described and is considered the standard for determination of neutralizing virus-specific antibody titers (Calisher et al., 1989; Lindsey et al., 1976; Shambaugh et al., 2017). For the detection of SARS-CoV-2 neutralizing antibodies, 2-fold serially diluted test serum (100μl) was mixed with 100μl of 150-200 plaque forming units (PFUs) of SARS-CoV-2 (isolate USA-WA1/2020, BEI Resources, NR-52281) and incubated for 1 h at 37°C, 5% CO_2_. The virus:serum mixture (100μl) was applied to VeroE6 cells (C1008, ATCC CRL-1586) grown to 95-100% confluency in 6 well plates. Adsorption of the virus:serum mixture was allowed to proceed for 1 hour at 37°C, 5% CO_2._ Following the adsorption period, a 0.6% agar overlay prepared in cell culture medium (Eagle’s Minimal Essential Medium, 2% heat inactivated FBS, 100μg/ml Penicillin G, 100 U/ml Streptomycin) was applied. Two days post-infection, a second agar overlay containing 0.2% neutral red prepared in cell culture medium was applied, and the number of plaques in each sample well were recorded after an additional 1-2 days incubation. Neutralizing titers were defined as the inverse of the highest dilution of serum providing 50% (PRNT50) or 90% (PRNT90) viral plaque reduction relative to a virus only control.

### Data Analysis

Subjects with known disease severity and sex were included in ordered probit regression analysis. The data was randomly separated into training (70%) and testing (30%) subsets. Data was then centered by subtracting the mean of each predictor from that predictor’s values, and then scaled by dividing by the standard deviation. Imputation of missing values was performed using the k-nearest neighbors algorithm. Each created model’s accuracy was assessed by measuring its performance on the testing subset of data. One-sided binomial tests were performed to determine if the accuracy of each model was significantly better than the no-information rate. Variable selection for the final model was performed by backwards stepwise selection by Akaike information criterion.

For construction of the correlation matrix, Spearman’s correlations were calculated using all complete pairs of variables in the dataset. All associated p-values were then adjusted via the Benjamini and Hochberg method. The network created using these Spearman’s correlations was gated to only include those relationships with an r_s_ > 0.65. Eigenvector centrality was calculated for each variable and is represented by the size of the respective node. Edge width corresponds with the strength of each correlation.

R version 4.0.2 was used for the probit regression modeling and construction of the correlation matrix (R Core Team (2020). R: A language and environment for statistical computing. R Foundation for Statistical Computing, Vienna, Austria. URL https://www.R-project.org/.). The caret, MASS, and RANN packages were also used for the probit regression (Max Kuhn (2020). caret: Classification and Regression Training. R package version 6.0-86. https://CRAN.R-project.org/package=caret)(Venables, W. N. & Ripley, B. D. (2002) Modern Applied Statistics with S. Fourth Edition. Springer, New York. ISBN 0-387-95457-0) (Sunil Arya, David Mount, Samuel E. Kemp and Gregory Jefferis (2019). RANN: Fast Nearest Neighbour Search (Wraps ANN Library) Using L2 Metric. R package version 2.6.1. https://CRAN.R-project.org/package=RANN). The corrplot package was used to create the correlation matrix (Taiyun Wei and Viliam Simko (2017). R package “corrplot”: Visualization of a Correlation Matrix (Version 0.84). Available from. https://github.com/taiyun/corrplot). Gephi 0.9.2 was used in the construction of the correlation network (Bastian M., Heymann S., Jacomy M. (2009). Gephi: an open source software for exploring and manipulating networks. International AAAI Conference on Weblogs and Social Media.).

---

**Supplemental Table 1:**

|  | IgM |  |  |  | IgA |  |  |  | IgG |  |  |  |
| --- | --- | --- | --- | --- | --- | --- | --- | --- | --- | --- | --- | --- |
|  | NP | RBD | S1 | S2 | NP | RBD | S1 | S2 | NP | RBD | S1 | S2 |
| Average Index Value | 1.35 | 6.45 | 9.75 | 0.50 | 1.51 | 3.86 | 6.06 | 3.14 | 8.49 | 12.43 | 19.68 | 23.21 |
| Positive | 22% | 73% | 79% | 5% | 22% | 60% | 70% | 34% | 97% | 97% | 98% | 98% |
| Negative | 78% | 27% | 21% | 95% | 78% | 40% | 30% | 66% | 3% | 3% | 2% | 2% |

**Supplemental Table 2:**

|  |  | Average Index Value | Positive | Negative |
| --- | --- | --- | --- | --- |
| IgG1 | NP | 19.03 | 97% | 3% |
|  | RBD | 18.71 | 91% | 9% |
|  | S1 | 137.97 | 97% | 3% |
|  | S2 | 16.43 | 53% | 47% |
| IgG2 | NP | 1.61 | 21% | 10% |
|  | RBD | 1.09 | 10% | 90% |
|  | S1 | 1.69 | 14% | 86% |
|  | S2 | 3.20 | 8% | 92% |
| IgG3 | NP | 4.93 | 64% | 36% |
|  | RBD | 3.91 | 39% | 61% |
|  | S1 | 6.18 | 59% | 41% |
|  | S2 | 43.90 | 94% | 6% |
| IgG4 | NP | 1.58 | 9% | 91% |
|  | RBD | 0.61 | 1% | 99% |
|  | S1 | 0.36 | 1% | 99% |
|  | S2 | 0.24 | 0% | 100% |

**Supplemental Table 3:**
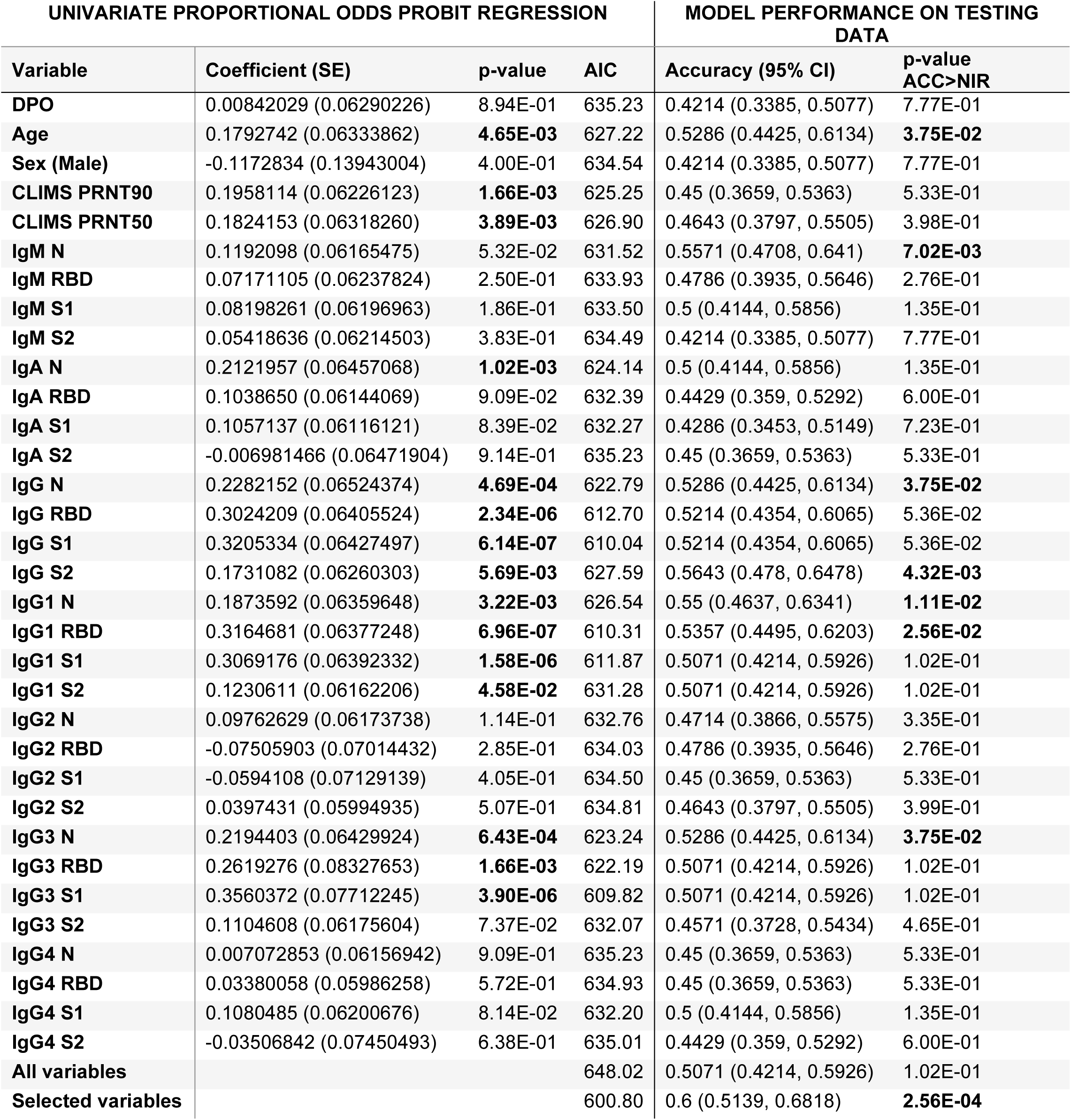

| UNIVARIATE PROPORTIONAL ODDS PROBIT REGRESSION |  |  |  | MODEL PERFORMANCE ON TESTING DATA |  |
| --- | --- | --- | --- | --- | --- |
| Variable | Coefficient (SE) | p-value | AIC | Accuracy (95% CI) | p-value<br>ACC>NIR |
| DPO | 0.00842029 (0.06290226) | 8.94E-01 | 635.23 | 0.4214 (0.3385, 0.5077) | 7.77E-01 |
| Age | 0.1792742 (0.06333862) | **4.65E-03** | 627.22 | 0.5286 (0.4425, 0.6134) | **3.75E-02** |
| Sex (Male) | -0.1172834 (0.13943004) | 4.00E-01 | 634.54 | 0.4214 (0.3385, 0.5077) | 7.77E-01 |
| CLIMS PRNT90 | 0.1958114 (0.06226123) | **1.66E-03** | 625.25 | 0.45 (0.3659, 0.5363) | 5.33E-01 |
| CLIMS PRNT50 | 0.1824153 (0.06318260) | **3.89E-03** | 626.90 | 0.4643 (0.3797, 0.5505) | 3.98E-01 |
| IgM N | 0.1192098 (0.06165475) | 5.32E-02 | 631.52 | 0.5571 (0.4708, 0.641) | **7.02E-03** |
| IgM RBD | 0.07171105 (0.06237824) | 2.50E-01 | 633.93 | 0.4786 (0.3935, 0.5646) | 2.76E-01 |
| IgM S1 | 0.08198261 (0.06196963) | 1.86E-01 | 633.50 | 0.5 (0.4144, 0.5856) | 1.35E-01 |
| IgM S2 | 0.05418636 (0.06214503) | 3.83E-01 | 634.49 | 0.4214 (0.3385, 0.5077) | 7.77E-01 |
| IgA N | 0.2121957 (0.06457068) | **1.02E-03** | 624.14 | 0.5 (0.4144, 0.5856) | 1.35E-01 |
| IgA RBD | 0.1038650 (0.06144069) | 9.09E-02 | 632.39 | 0.4429 (0.359, 0.5292) | 6.00E-01 |
| IgA S1 | 0.1057137 (0.06116121) | 8.39E-02 | 632.27 | 0.4286 (0.3453, 0.5149) | 7.23E-01 |
| IgA S2 | -0.006981466 (0.06471904) | 9.14E-01 | 635.23 | 0.45 (0.3659, 0.5363) | 5.33E-01 |
| IgG N | 0.2282152 (0.06524374) | **4.69E-04** | 622.79 | 0.5286 (0.4425, 0.6134) | **3.75E-02** |
| IgG RBD | 0.3024209 (0.06405524) | **2.34E-06** | 612.70 | 0.5214 (0.4354, 0.6065) | 5.36E-02 |
| IgG S1 | 0.3205334 (0.06427497) | **6.14E-07** | 610.04 | 0.5214 (0.4354, 0.6065) | 5.36E-02 |
| IgG S2 | 0.1731082 (0.06260303) | **5.69E-03** | 627.59 | 0.5643 (0.478, 0.6478) | **4.32E-03** |
| IgG1 N | 0.1873592 (0.06359648) | **3.22E-03** | 626.54 | 0.55 (0.4637, 0.6341) | **1.11E-02** |
| IgG1 RBD | 0.3164681 (0.06377248) | **6.96E-07** | 610.31 | 0.5357 (0.4495, 0.6203) | **2.56E-02** |
| IgG1 S1 | 0.3069176 (0.06392332) | **1.58E-06** | 611.87 | 0.5071 (0.4214, 0.5926) | 1.02E-01 |
| IgG1 S2 | 0.1230611 (0.06162206) | **4.58E-02** | 631.28 | 0.5071 (0.4214, 0.5926) | 1.02E-01 |
| IgG2 N | 0.09762629 (0.06173738) | 1.14E-01 | 632.76 | 0.4714 (0.3866, 0.5575) | 3.35E-01 |
| IgG2 RBD | -0.07505903 (0.07014432) | 2.85E-01 | 634.03 | 0.4786 (0.3935, 0.5646) | 2.76E-01 |
| IgG2 S1 | -0.0594108 (0.07129139) | 4.05E-01 | 634.50 | 0.45 (0.3659, 0.5363) | 5.33E-01 |
| IgG2 S2 | 0.0397431 (0.05994935) | 5.07E-01 | 634.81 | 0.4643 (0.3797, 0.5505) | 3.99E-01 |
| IgG3 N | 0.2194403 (0.06429924) | **6.43E-04** | 623.24 | 0.5286 (0.4425, 0.6134) | **3.75E-02** |
| IgG3 RBD | 0.2619276 (0.08327653) | **1.66E-03** | 622.19 | 0.5071 (0.4214, 0.5926) | 1.02E-01 |
| IgG3 S1 | 0.3560372 (0.07712245) | **3.90E-06** | 609.82 | 0.5071 (0.4214, 0.5926) | 1.02E-01 |
| IgG3 S2 | 0.1104608 (0.06175604) | 7.37E-02 | 632.07 | 0.4571 (0.3728, 0.5434) | 4.65E-01 |
| IgG4 N | 0.007072853 (0.06156942) | 9.09E-01 | 635.23 | 0.45 (0.3659, 0.5363) | 5.33E-01 |
| IgG4 RBD | 0.03380058 (0.05986258) | 5.72E-01 | 634.93 | 0.45 (0.3659, 0.5363) | 5.33E-01 |
| IgG4 S1 | 0.1080485 (0.06200676) | 8.14E-02 | 632.20 | 0.5 (0.4144, 0.5856) | 1.35E-01 |
| IgG4 S2 | -0.03506842 (0.07450493) | 6.38E-01 | 635.01 | 0.4429 (0.359, 0.5292) | 6.00E-01 |
| All variables |  |  | 648.02 | 0.5071 (0.4214, 0.5926) | 1.02E-01 |
| Selected variables |  |  | 600.80 | 0.6 (0.5139, 0.6818) | **2.56E-04** |

**Supplemental Table 4:**

| PROPORTIONAL ODDS PROBIT REGRESSION WITH AGE AS A COVARIATE |  |  |  | MODEL PERFORMANCE ON TESTING DATA |  |
| --- | --- | --- | --- | --- | --- |
| Variable | Coefficient (SE) | p-value | AIC | Accuracy (95% CI) | p-value ACC>NIR |
| DPO | -0.004119273 (0.06338651) | 9.48E-01 | 629.22 | 0.5286 (0.4425, 0.6134) | **3.75E-02** |
| Sex (Male) | -0.1293131 (0.13987964) | 3.55E-01 | 628.36 | 0.5429 (0.4566, 0.6272) | **1.71E-02** |
| CLIMS PRNT90 | 0.1559160 (0.06547446) | **1.73E-02** | 623.51 | 0.5429 (0.4566, 0.6272) | **1.71E-02** |
| CLIMS PRNT50 | 0.1449436 (0.06572414) | **2.74E-02** | 624.36 | 0.5214 (0.4354, 0.6065) | 5.36E-02 |
| IgM N | 0.07836274 (0.06409214) | 2.21E-01 | 627.73 | 0.5214 (0.4354, 0.6065) | 5.36E-02 |
| IgM RBD | 0.01943351 (0.06557456) | 7.67E-01 | 629.13 | 0.55 (0.4637, 0.6341) | **1.11E-02** |
| IgM S1 | 0.02322937 (0.06622030) | 7.26E-01 | 629.10 | 0.5571 (0.4708, 0.641) | **7.02E-03** |
| IgM S2 | 0.05514419 (0.06216877) | 3.75E-01 | 628.44 | 0.5643 (0.478, 0.6478) | **4.32E-03** |
| IgA N | 0.1710039 (0.06742154) | **1.12E-02** | 622.63 | 0.5429 (0.4566, 0.6272) | **1.71E-02** |
| IgA RBD | 0.09119512 (0.06167864) | 1.39E-01 | 627.04 | 0.5357 (0.4495, 0.6203) | **2.56E-02** |
| IgA S1 | 0.09161639 (0.06145615) | 1.36E-01 | 627.01 | 0.5143 (0.4284, 0.5996) | 7.47E-02 |
| IgA S2 | -0.0227294 (0.06522780) | 7.27E-01 | 629.10 | 0.5214 (0.4354, 0.6065) | 5.36E-02 |
| IgG N | 0.1942685 (0.06750208) | **4.00E-03** | 620.84 | 0.5571 (0.4708, 0.641) | **7.02E-03** |
| IgG RBD | 0.2820277 (0.06481207) | **1.35E-05** | 610.10 | 0.5929 (0.5067, 0.675) | **4.76E-04** |
| IgG S1 | 0.2993826 (0.06512311) | **4.28E-06** | 607.84 | 0.5357 (0.4495, 0.6203) | **2.56E-02** |
| IgG S2 | 0.1487403 (0.06345589) | **1.91E-02** | 623.72 | 0.5857 (0.4995, 0.6683) | **8.61E-04** |
| IgG1 N | 0.1443364 (0.06732723) | **3.20E-02** | 624.62 | 0.5929 (0.5067, 0.675) | **4.76E-04** |
| IgG1 RBD | 0.2937825 (0.06482250) | **5.84E-06** | 608.46 | 0.5786 (0.4923, 0.6615) | **1.52E-03** |
| IgG1 S1 | 0.2837305 (0.06491573) | **1.24E-05** | 609.88 | 0.5214 (0.4354, 0.6065) | 5.36E-02 |
| IgG1 S2 | 0.09804867 (0.06240420) | 1.16E-01 | 626.76 | 0.5571 (0.4708, 0.641) | **7.02E-03** |
| IgG2 N | 0.08248615 (0.06185257) | 1.82E-01 | 627.45 | 0.5286 (0.4425, 0.6134) | **3.75E-02** |
| IgG2 RBD | -0.06202069 (0.07071954) | 3.80E-01 | 628.41 | 0.5357 (0.4495, 0.6203) | **2.57E-02** |
| IgG2 S1 | -0.05381328 (0.07181056) | 4.54E-01 | 628.62 | 0.5357 (0.4495, 0.6203) | **2.57E-02** |
| IgG2 S2 | 0.03299862 (0.06017975) | 5.83E-01 | 628.92 | 0.5357 (0.4495, 0.6203) | **2.57E-02** |
| IgG3 N | 0.1925910 (0.06495653) | **3.03E-03** | 620.21 | 0.5714 (0.4851, 0.6547) | **2.59E-03** |
| IgG3 RBD | 0.2625168 (0.08549222) | **2.14E-03** | 616.70 | 0.5 (0.4144, 0.5856) | 1.35E-01 |
| IgG3 S1 | 0.3463172 (0.07767068) | **8.24E-06** | 605.61 | 0.5429 (0.4566, 0.6272) | **1.71E-02** |
| IgG3 S2 | 0.1169920 (0.06213372) | 5.97E-02 | 625.71 | 0.5 (0.4144, 0.5856) | 1.35E-01 |
| IgG4 N | -0.008487359 (0.06277202) | 8.92E-01 | 629.20 | 0.5286 (0.4425, 0.6134) | **3.75E-02** |
| IgG4 RBD | 0.04314625 (0.06013673) | 4.73E-01 | 628.70 | 0.5286 (0.4425, 0.6134) | **3.75E-02** |
| IgG4 S1 | 0.1071146 (0.06217504) | 8.49E-02 | 626.25 | 0.5286 (0.4425, 0.6134) | **3.75E-02** |
| IgG4 S2 | -0.02925658 (0.07612034) | 7.01E-01 | 629.06 | 0.5286 (0.4425, 0.6134) | **3.75E-02** |

**Supplemental Figure 1:**
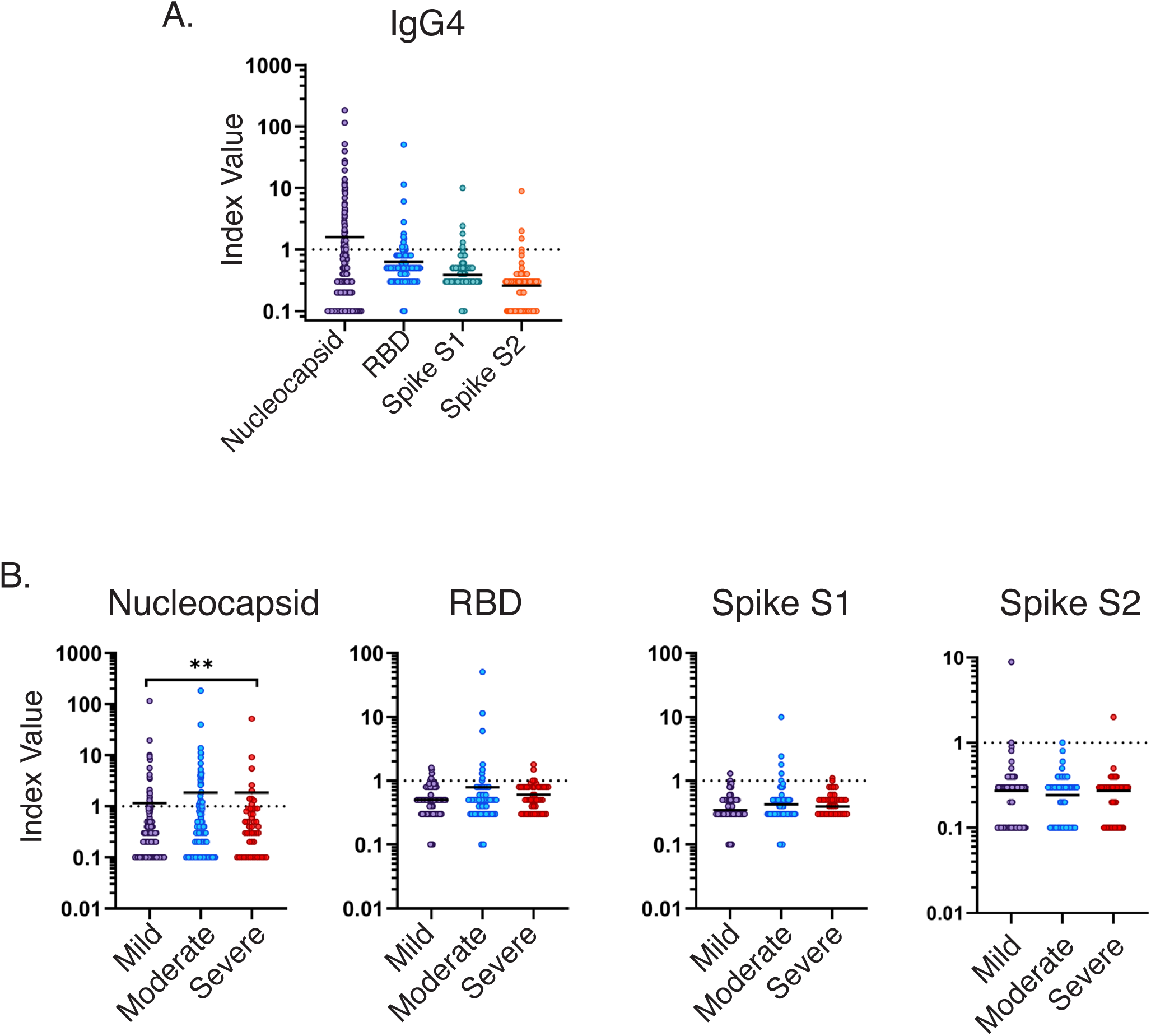
IgG4

**Supplemental Figure 2:**
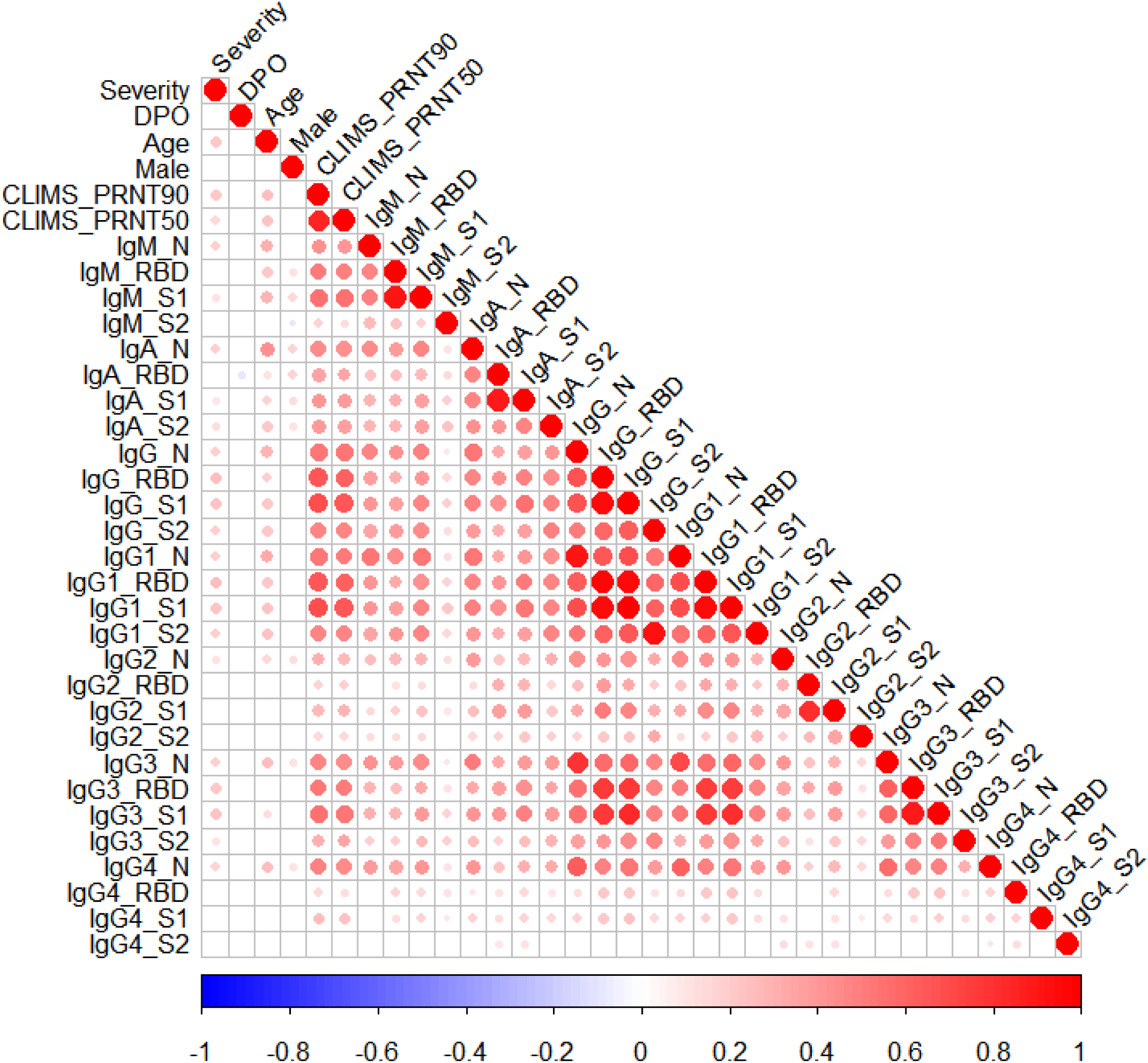
Correlation Matrix

